# Kidney ‘pathway-orphan’ genes as a potential overlooked source of novel insights into kidney health and disease

**DOI:** 10.1101/2024.07.09.24309717

**Authors:** Dianne Acoba, Sophie Gunnarsson, Anna Reznichenko

## Abstract

Hundreds of protein-coding genes still have unknown functions and no biological pathway annotations. Mechanistic studies typically investigate well-known genes, despite growing evidence of associations between disease and some understudied genes. We hypothesized that examining these overlooked genes lacking pathway annotation could lead to new insights on chronic kidney disease (CKD) pathogenesis. Human protein-coding genes (N=19,258) from HGNC were annotated with pathway membership using a comprehensive collection of pathway databases (including but not limited to GO, KEGG, MSigDB, Reactome, WikiPathways) to reveal ‘pathway-orphan genes’— genes that are not members of any biological pathways. Expression and enrichment of pathway-orphan genes in healthy kidneys were established using GTEx data. Kidney-expressed pathway-orphan genes were tested for differential case-control expression using publicly available CKD datasets with kidney tissue RNA-seq transcriptomics profiling (GSE98422, GSE142025, GSE175759, GSE197307, Levin et al., 2020). A total of 286 genes in the human genome currently lack any biological pathway membership and are identified as pathway-orphan genes. We have determined 97 of these pathway-orphan genes are expressed in healthy kidneys, with several showing kidney-specific enrichment. Furthermore, 34 pathway-orphan genes show significant modulation of expression (FDR < 0.05) in CKD kidney, out of which 10 robustly demonstrate concordant directionality of change in more than one RNA-seq study. Through interrogating multiple lines of evidence, we showed how possible physiological functions of the pathway-orphan genes in kidney health and disease can be inferred. A substantial number of kidney-expressed genes remain ‘pathway-orphan’ while displaying clear signals of kidney relevance, such as enrichment of expression in the kidney, significant modulation in CKD, and genetic associations with kidney function. Directing mechanistic studies into this overlooked gene group might broaden our biological understanding of kidney physiology and highlight novel disease drivers.

## INTRODUCTION

Giving biological pathway context is paramount to biomedical research but pathway analysis is limited by the pre-existing pathway entries in annotation databases, such as Gene Ontology (GO), Reactome, and Kyoto Encyclopedia of Genes and Genomes (KEGG) [1]. Despite the human genome being published more than 20 years ago, there are still protein-coding genes with no pathway membership in annotation databases as functional studies typically focus on well-studied genes [2]. Such inequality in research attention gets amplified with time as the understudied genes remain overlooked and the ‘rich get richer’ with pathway and functional annotations [1].

This attention bias is commonplace in biological research and is also exhibited by publication trends—95% of all life science publications focus on only 5 000 human proteins [1–4]. Factors perpetuating this inequality are limited prior knowledge, biased and unequal annotation databases, reagent availability, funding granted to projects with preceding evidence, risk-averse researchers, preference for experimentally well-accessible genes, and a shortage of large-scale perturbation studies [3, 5].

Disease understanding and drug discovery for chronic kidney disease (CKD) has been historically difficult due to its heterogeneous etiologies, which may involve more than 100 pathways in different crosstalk architectures across diagnoses [6, 7]. This molecular complexity, however, while a challenge, also allows the use of high-throughput or omics data from large patient cohorts to extensively identify potential disease drivers and candidate drug targets [6, 8]. These omics studies allow researchers to form hypotheses for follow-up mechanistic studies, by triaging and prioritizing genes with available evidence of their possible pathophysiological role [1].

In this study, we hence set out to systematically identify and investigate protein-coding genes without any known biological pathway annotations in the kidneys of both healthy individuals and CKD patients. We *in silico* characterize and compare these ‘pathway-orphan’ genes to ‘pathway-annotated’ and housekeeping genes. Our hypothesis is that examining this previously overlooked group of genes may lead to new insights into the disease pathogenesis of CKD.

## MATERIALS AND METHODS

All databases were accessed on 26 January 2024, and the versions utilized are provided in Supplementary Table 1.

A list of protein-coding genes (N=19,258) was downloaded from the HUGO Gene Nomenclature Committee (HGNC) and annotated using a comprehensive collection of pathway databases: Gene Ontology (GO) Biological Process (BP) and Molecular Function (MF) terms, Reactome, Kyoto Encyclopedia of Genes and Genomes (KEGG), Hallmark and Curated gene sets from Molecular Signatures Database (MSigDB), WikiPathways, Ingenuity Pathway Analysis (IPA), and 28 other pathway repositories in ConsensusPathDB (summarized in **Supplementary Table 2**).

Gene length information was extracted from GENCODE, while protein information (length, mass, domains and families) were downloaded from UniProt and InterPro. DeepLoc 2.0 was additionally used to predict subcellular location. The counts of gene-associated publications indexed on PubMed were extracted using the *gene2pubmed* file obtained from the NCBI website (https://ftp.ncbi.nlm.nih.gov/gene/DATA/gene2pubmed.gz).

Expression levels of the protein-coding genome in healthy kidneys were established using RNA-seq data retrieved from Genotype-Tissue Expression (GTEx) v8 and Human Protein Atlas (HPA) v23. The presence of detectable gene expression in the kidney was defined as TPM > 0 in at least 75% of GTEx samples (63 out of 85 cortex samples and 3 out of 4 medulla samples). To compute a measure of kidney-specific enrichment of expression as compared to other tissues, the per-tissue mean expression for each gene was subjected to Z-transformation across tissues and then to a second Z-transformation across genes to bring all Z-scores to the same scale.

A list of 407 housekeeping genes (genes ubiquitously expressed in all tissue and cell types) was extracted from literature [9]. Pathway-annotated genes were defined as protein-coding genes that are neither pathway-orphan nor housekeeping genes. A list of genes with protein products enriched in the kidney was also extracted from HPA. Modulation of the expression in disease was tested using publicly available CKD kidney tissue RNA-seq transcriptomics datasets (GSE142025, GSE175759, GSE197307, GSE98422, Levin et al [10]). DESeq2 was used to perform differential expression analysis contrasting CKD vs control, and genes with false discovery rate (FDR) < 0.05 were considered differentially expressed [11]. Pathway-orphan genes with case-control differential expression in at least two CKD studies were further *in silico* characterized.

Expression during fetal kidney development was determined using the DESCARTES and Bgee databases and a study by Lindström et al [12]. GWAS Catalog was used to query for genetic associations, HumanBase for predicted functions, BioGrid for known interactors, Alliance of Genome Resources (AGR) for orthologs in other species, and Pharos for other predicted annotations. Nephroseq v5 was used to check for pathway-orphan differential expression and gene expression correlation with clinical variables, such as glomerular filtration rate (GFR), proteinuria, and blood urea nitrogen (BUN). GeneHancer was used to determine if pathway-orphan genes have regulatory enhancers with single-nucleotide polymorphisms (SNPs) that affect kidney function phenotypes. NephQTL2 and GTEx were queried to check for cis-eQTLs present within 1 megabase pairs of the pathway-orphan genes in the kidney.

All analyses were performed in R v4.1.3 [13].

## RESULTS

### Pathway-orphan genes identification and general characteristics

Following our analysis strategy (**Figure 1**), we reveal that almost two percent of the protein-coding human genome—286 genes—do not belong to any of the currently known pathways or gene sets. We now refer to these unannotated genes as ‘pathway-orphan genes’.

**Figure 1.**
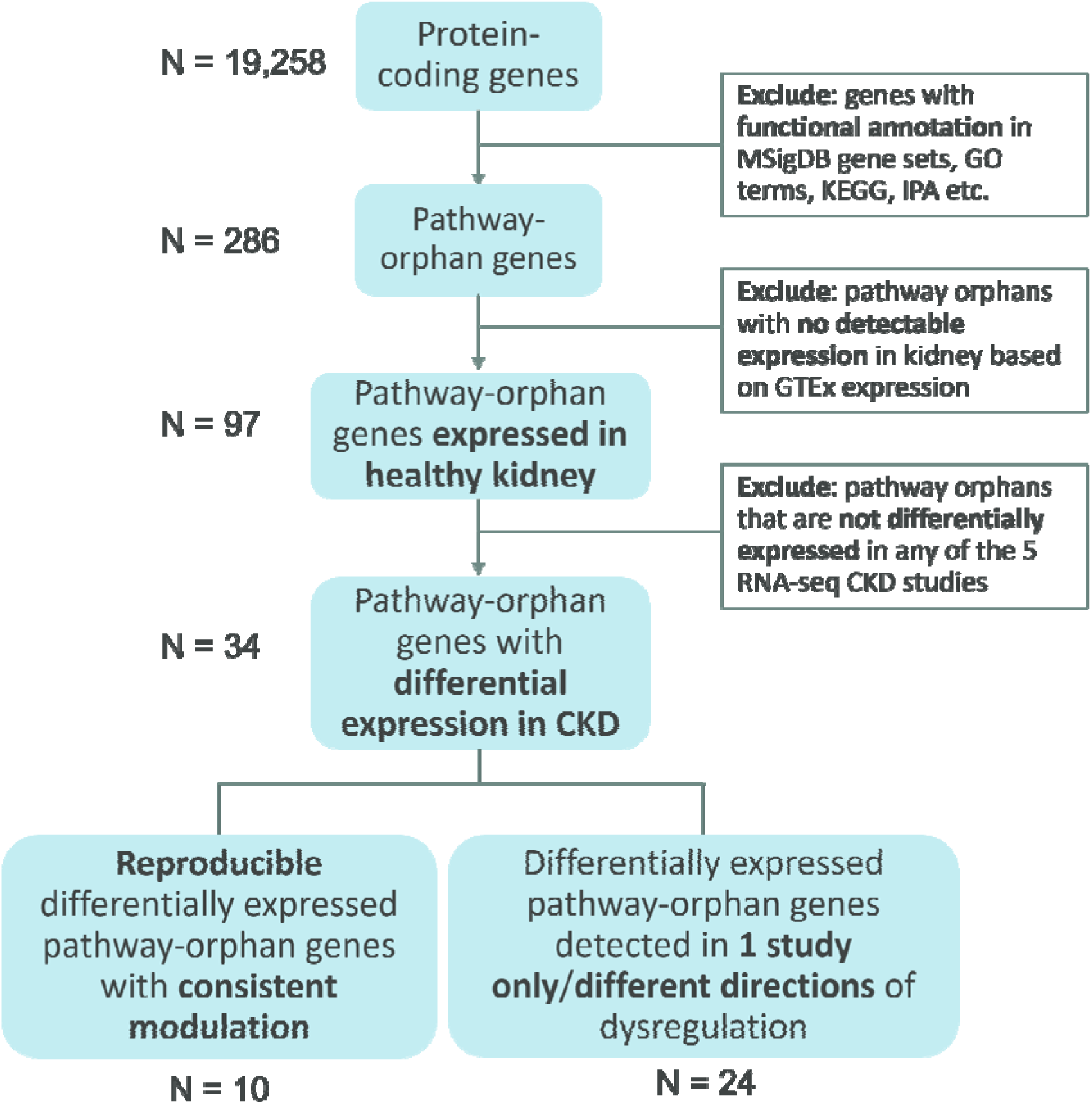
Analysis scheme. The flowchart shows the stepwise process of pathway-orphan genes identification and annotation in the chronic kidney disease context.

Pathway-orphan genes are distributed across all 24 human chromosomes, with the X chromosome harboring the highest number comprising over 6% of its total gene count (**Figure 2A**). As expected, pathway-orphan genes have substantially fewer associated publications compared to both pathway-annotated and housekeeping genes, with 17% having zero publications on PubMed (covering all articles indexed on PubMed) (**Figure 2B**).

**Figure 2.**
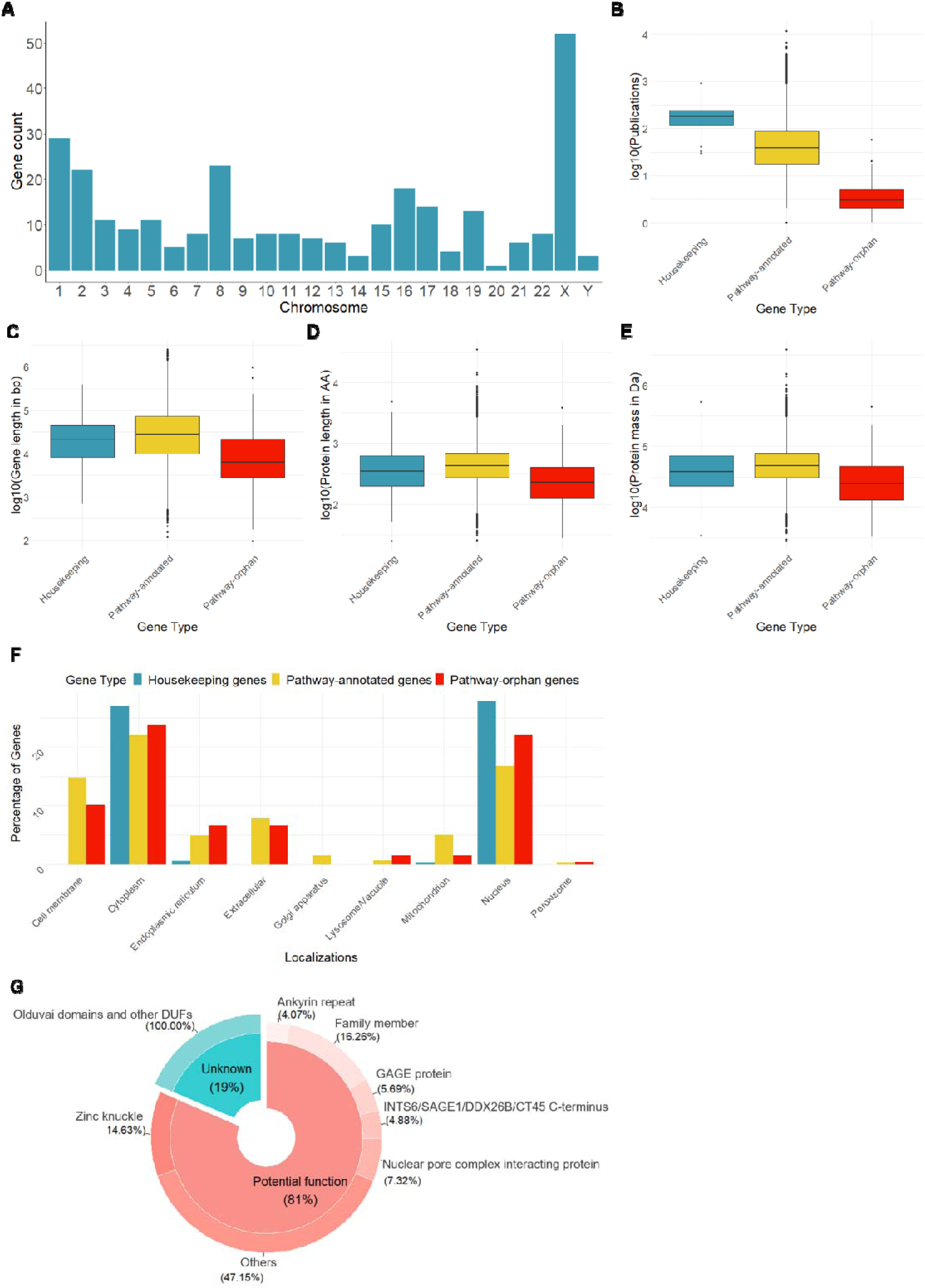
Pathway-orphan genes characteristics and trends. (A) Chromosomal location of pathway-orphan genes. The barplot shows the pathway-orphan genes distribution across all 24 chromosomes, as reported by HGNC. The x-axis corresponds to the chromosome number and the y-axis to the pathway-orphan gene count. (B) Boxplots show the values and distribution of the number of publications indexed on PubMed of the three protein-coding gene types. The y-axis corresponds to the log10-transformed publication count. (C) Boxplots show the values and distribution of the gene lengths of the three protein-coding gene types. The y-axis corresponds to the log10-transformed gene length in base pair units. (D) Boxplots show the values and distribution of the protein lengths of the three protein-coding gene types. The y-axis corresponds to the log10-transformed protein length in number of amino acids. (E) Boxplots show the values and distribution of the protein mass of the three protein-coding gene types. The y-axis corresponds to the log10-transformed protein mass in Daltons. (F) The barplot shows how the different protein-coding gene types are localized in each subcellular compartment percentage-wise over total gene count of each type. (G) Donut chart shows the most common domains in pathway-orphan proteins. DUF: Domain of Unknown Functions.

With respect to gene length, pathway-orphan genes are generally significantly shorter, with mean length of 23.82 kilobase pairs (kbp), compared to housekeeping genes (mean: 39.10 kbp, *P* < 0.001) and pathway-annotated genes (mean: 69.92 kbp, *P* < 0.001). Consequently, the same tendency is observed in protein length where the mean length of pathway-orphan proteins is 320 amino acids long, compared to 514 (*P* < 0.001) and 582 (*P* < 0.001) for housekeeping and pathway-annotated proteins, respectively. Pathway-orphan proteins are also lighter in molecular weight, with mean mass of 35.61 kilodaltons (kDa), whereas housekeeping proteins have a mean of 57.50 kDa (*P* < 0.001) and 64.77 kDa (*P* < 0.001) for pathway-annotated proteins. **Figures 2C-E** show the value distribution of these attributes.

For subcellular localization of the expressed proteins, pathway-orphan genes are predicted to be predominantly cytoplasmic (N = 110) and nuclear localization comes in close second (N = 101). Distributions of protein localization for the three classes of protein-coding genes are similar and are shown in **Figure 2F**. Some genes are assumed to be present in more than one subcellular compartment and the localization breakdown is in **Supplementary Table 3**. Due to the limitations of DeepLoc, 120 protein-coding genes have no available subcellular localization prediction (26 pathway-orphan genes and 94 pathway-annotated genes).

Functional domain prediction could be accomplished for 151 out of the 286 pathway-orphan proteins. Out of these, 123 have at least one identified domain, while 28 have domains of unknown functions (**Figure 2G**). A full summary of the domain predictions of the pathway-orphan proteins is presented in **Supplementary Table 4**.

### Pathway-orphan genes in the healthy kidney

Analysis of the reference kidney expression data from GTEx reveals that 97 pathway-orphan genes have detectable expression levels in the kidney tissue, 80 in the cortex and 92 in the medulla, with an overlap of 75. **Supplementary Table 5** provides transcript expression and protein information on the 97 kidney-expressed pathway-orphan genes.

Kidney-expressed pathway-orphan genes retain the same characteristics of pathway-orphan genes in general, being significantly shorter in gene and protein length, with lighter protein molecular weight, predominantly cytoplasmic localization, and similar trends for predicted protein domains and families. Interestingly, pathway-orphan genes in the kidney have significantly lower mRNA expression levels as compared to pathway-annotated or housekeeping genes both in the cortex (**Figure 3A**) and medulla (**Figure 3B**). Of note, five pathway-orphan genes show specific enrichment of expression (specificity z-score > 2) [14] in the kidney cortex (*LY6L*, *C10orf106*, *MYOCOS*, *FAM240A*, and *SEC14L6*) and six in the medulla (*ERVV-2*, *FRG2C*, *MYOCOS*, *SEC14L6*, *FAM240A*, and *MED14OS*) as compared to other tissues and organs. **Figures 3C-D** show the distribution of tissue-specific enrichment, with known kidney-enriched genes as positive control.

**Figure 3.**
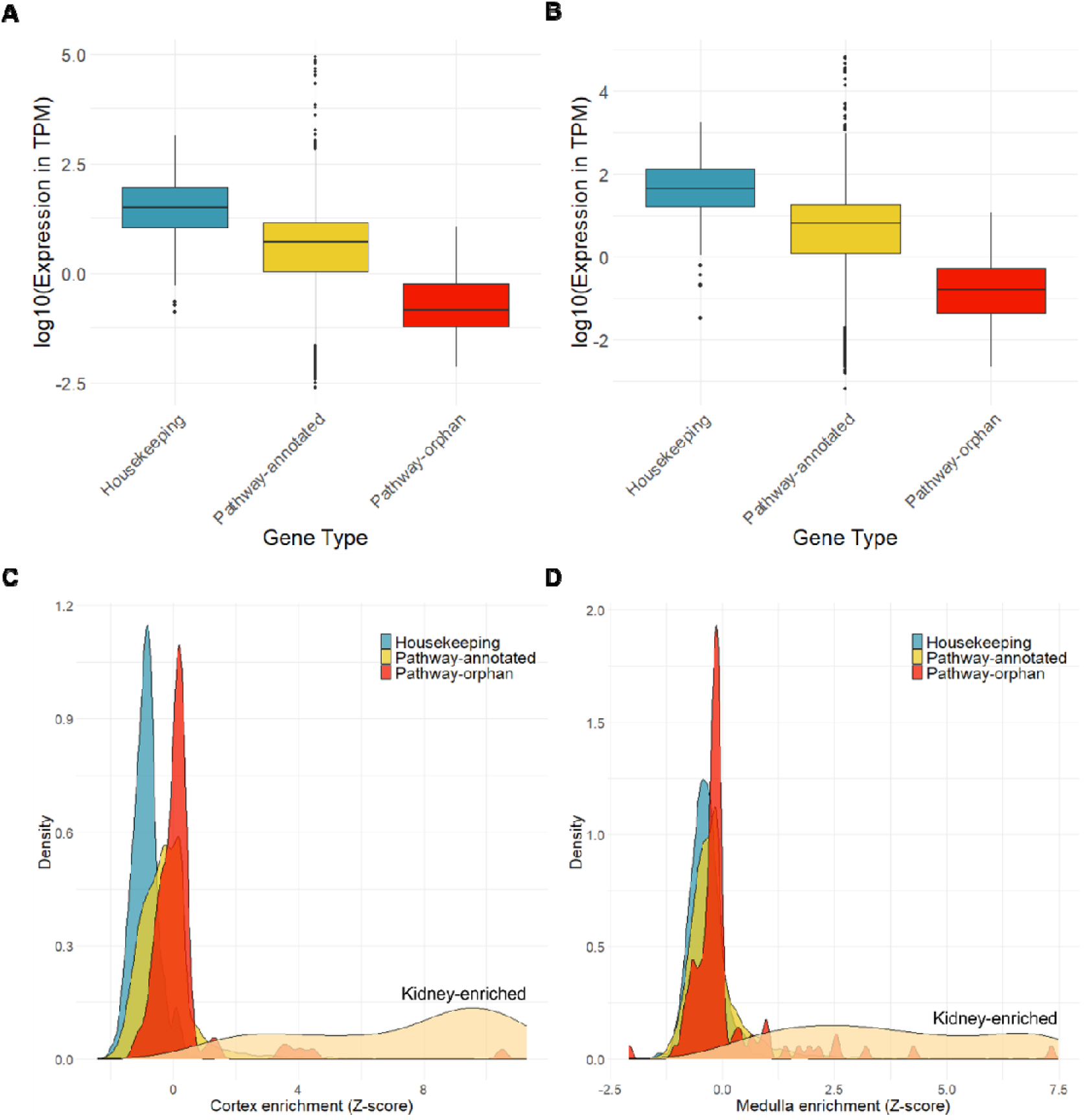
Kidney-expressed pathway-orphan genes characteristics and trends. (A, B) Boxplots show the values and distribution of the kidney gene expression values from GTEx of the three protein-coding gene types in the (A) cortex and (B) medulla. The y-axis corresponds to the log10-transformed transcript expression in TPM or transcripts-per-million. (C,D) Density plots show the distribution of the (C) cortex and (D) medulla enrichment z-scores calculated from GTEx of the three protein-coding gene types with kidney-enriched transcripts as positive control. The Z-scores of kidney-enriched genes extracted from the Human Protein Atlas were also plotted as positive control.

### Pathway-orphan genes modulation in CKD

Analysis of RNA-seq data from five independent studies identifies 34 pathway-orphan genes differentially expressed in the kidney tissues of CKD patients compared to healthy controls (**Figure 4**). Out of these, 10 pathway-orphan genes show concordant modulation directionality in two or more studies, 6 of which are downregulated (*C17orf107*, *C22orf31*, *C2CD4D*, *FAM229A*, *MED14OS*, and *STPG2*) and 4 that are upregulated (*C10orf105*, *IQANK1*, *NBPF26*, and *SEC14L6*) in disease vs control. In a dataset with micro-dissected kidney tissues, *C22orf31*, *FAM229A*, and *STPG2* are downregulated and *SEC14L6* is upregulated in both glomeruli and tubulointerstitium of CKD patients. Focusing on the 10 pathway-orphan genes that have concordant modulation in multiple studies, **Table 1** summarizes the evidence on the potential role of these pathway in CKD pathogenesis. **Table 2** shows differential expression in CKD and kidney measures correlation evidence extracted from Nephroseq.

**Figure 4.**
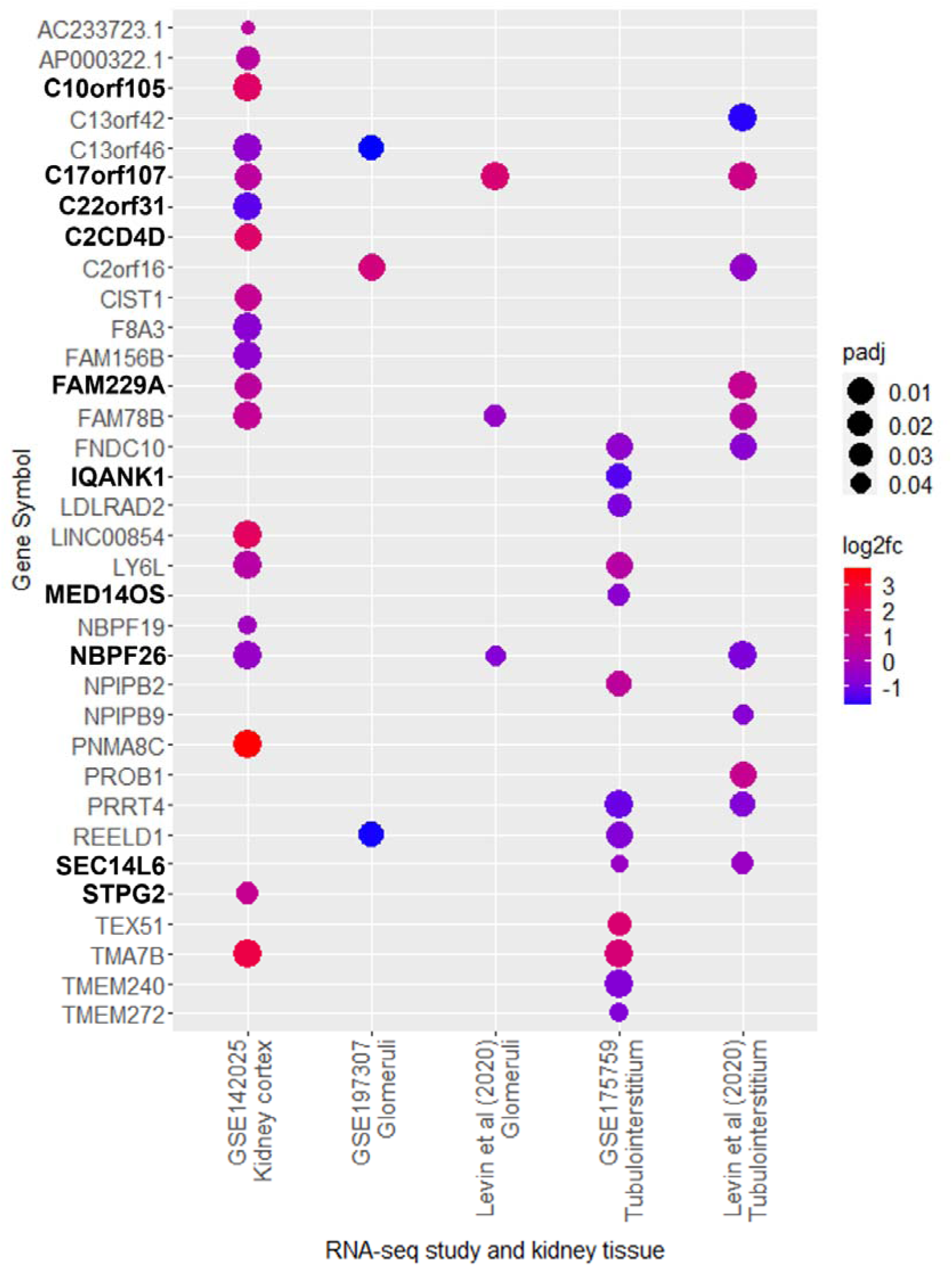
Dysregulation of pathway-orphan genes in chronic kidney disease. Dotplot shows the differentially expressed pathway-orphan genes when comparing diseased and healthy control kidney tissues. Only statistically significant values are shown. Size of the dots reflects the significance level, color indicates directionality of change. Genes in bold are the pathway-orphan genes with case-control differential expression in two or more studies.

**Table 1.**
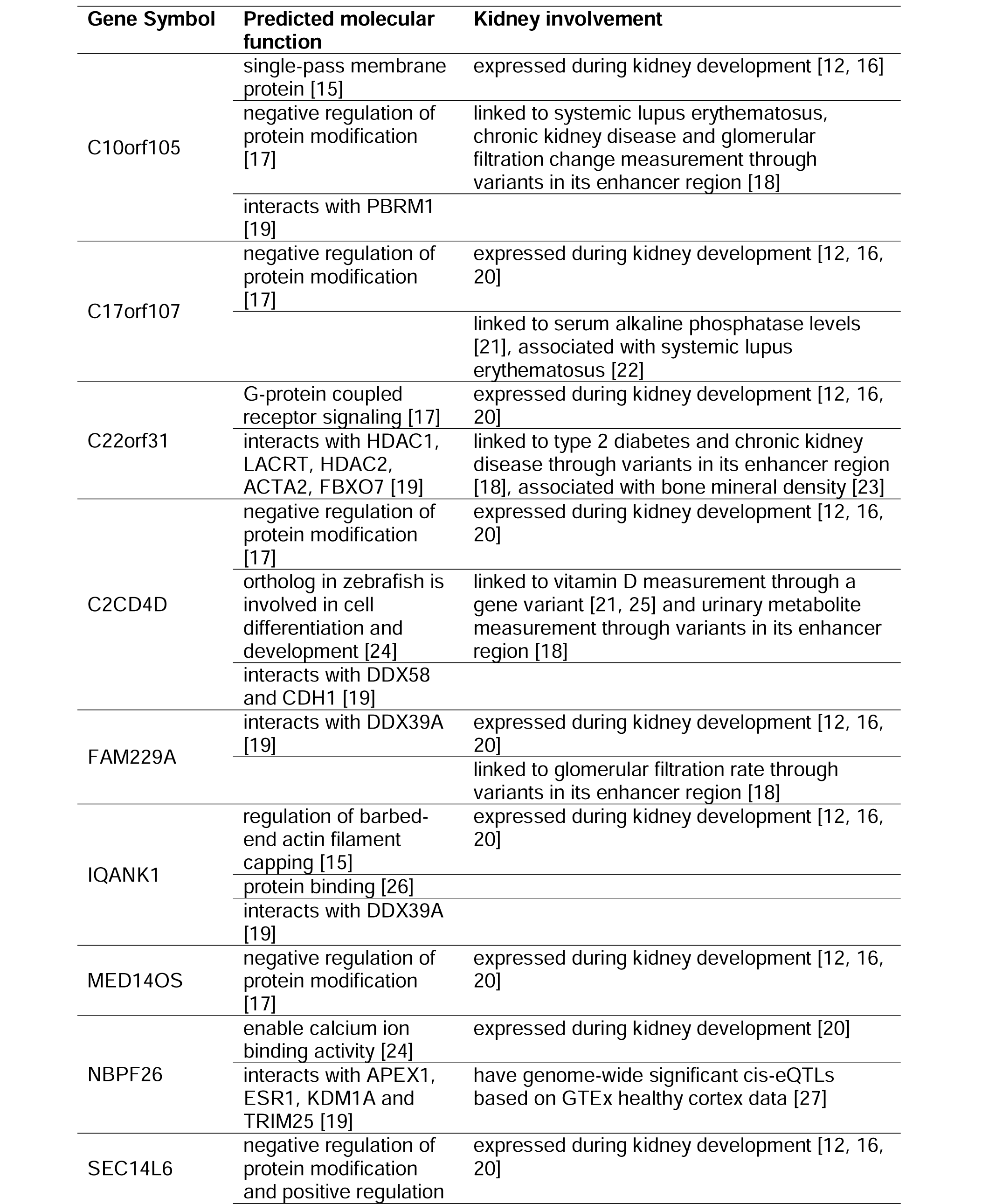
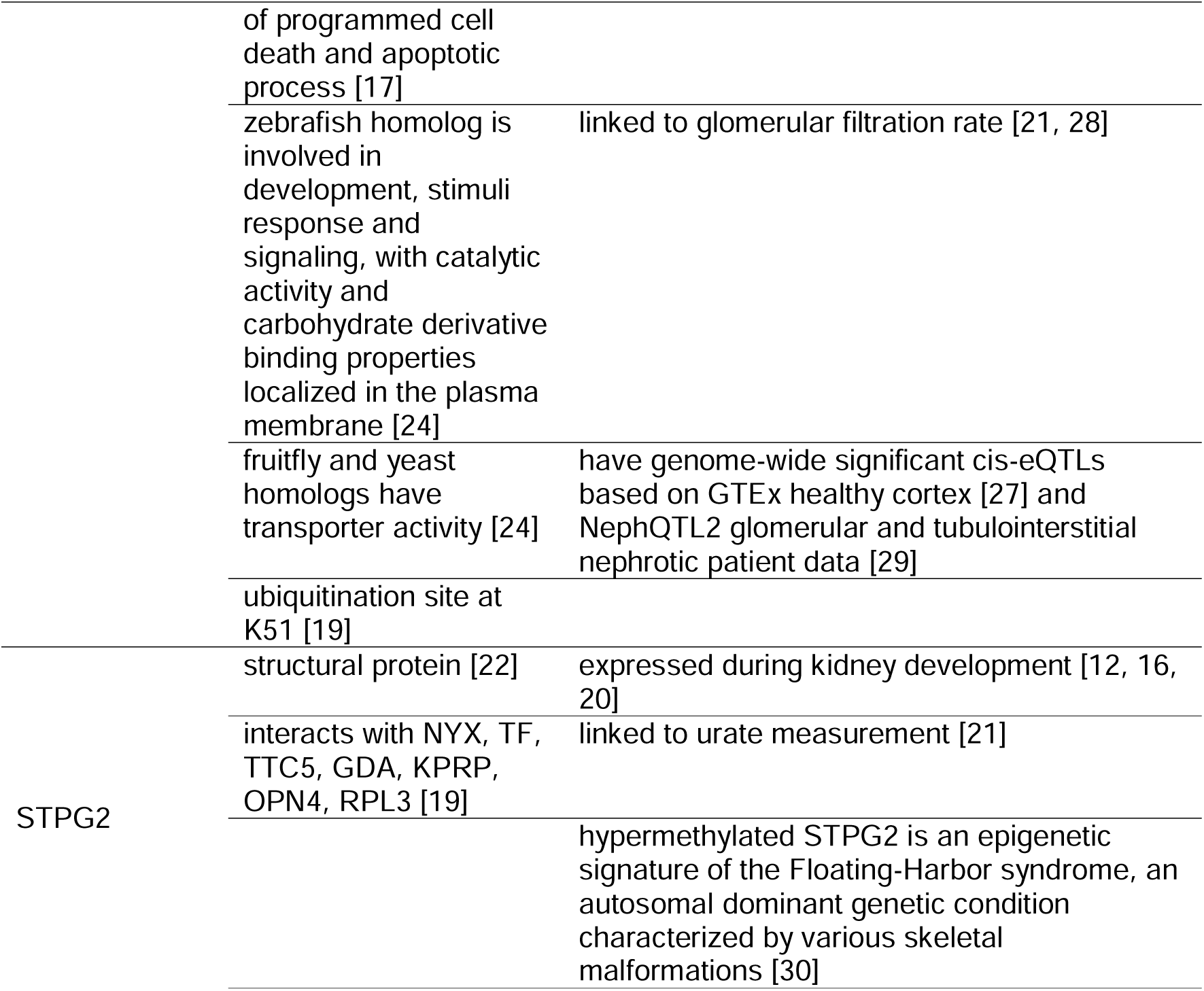
Summary of kidney-relevant evidence for the pathway-orphan genes robustly modulated in CKD.

**Table 2.** Compiled *in silico* evidence of pathway-orphan genes in CKD from Nephroseq.

| Gene | Finding | Effect size | P-value | Sample size | Dataset |
| --- | --- | --- | --- | --- | --- |
| C10orf105 | higher glomerular expression in FSGS patients with nephrotic and subnephrotic proteinuria vs normal proteinuria | 1.072* | 6.97E-7 | 91 | [31] |
|  | negative correlation of glomerular expression with baseline eGFR | -0.465<sup>†</sup> | 0.004 | 37 | [32] |
|  | lower tubulointerstitial expression in patients with *APOL1* high-risk genotype vs low-risk patients | -1.628* | 0.008 | 8 | [32] |
| C17orf107 | lower glomerular expression in patients with *APOL1* high-risk genotype vs low-risk patients | -1.279* | 0.018 | 38 | [32] |
|  | higher glomerular expression in FSGS patients with nephrotic proteinuria vs subnephrotic FSGS patients | 1.452* | 0.024 | 10 | [31] |
|  | negative correlation of tubulointerstitial expression with baseline BUN in LN patients | -0.787<sup>†</sup> | 0.036 | 7 | ERCB |
|  | positive correlation of glomerular expression with baseline proteinuria in MCD patients | 0.226<sup>†</sup> | 0.043 | 80 | [31] |
| C22orf31 | lower renal expression in CKD patients vs healthy controls | -2.166* | 5.74E-5 | 53 | [33] |
|  | lower tubulointerstitial expression in patients with DKD vs healthy controls | -1.282* | 0.002 | 22 | [34] |
|  | lower glomerular expression in FSGS patients vs healthy controls | -1.137* | 0.002 | 46 | [35] |
|  | lower tubulointerstitial expression in vasculitis patients vs healthy controls | -1.160* | 0.002 | 52 | [36] |
|  | lower tubulointerstitial expression in FSGS patients vs healthy controls | -1.131* | 0.004 | 48 | [36] |
|  | lower glomerular expression in LN patients vs healthy controls | -1.100* | 0.010 | 53 | [35] |
|  | lower glomerular expression in IgAN patients vs healthy controls | -1.085* | 0.027 | 48 | [35] |
|  | lower tubulointerstitial expression in LN patients vs healthy controls | -1.096* | 0.027 | 63 | [36] |
|  | lower glomerular expression in DKD patients vs healthy controls | -1.139* | 0.036 | 33 | [35] |
|  | lower tubulointerstitial expression in IgAN patients vs healthy controls | -1.090* | 0.047 | 56 | [36] |
|  | positive correlation of tubulointerstitial expression with eGFR | 0.237<sup>†</sup> | 0.001 | 186 | [36] |
|  | positive correlation of tubulointerstitial expression with baseline eGFR | 0.433<sup>†</sup> | 0.002 | 49 | [32] |
|  | positive correlation of glomerular expression with eGFR | 0.200<sup>†</sup> | 0.005 | 192 | [35] |
|  | positive correlation of glomerular expression with baseline eGFR in FSGS patients | 0.232<sup>†</sup> | 0.032 | 86 | [31] |
|  | positive correlation of glomerular expression with baseline eGFR in MCD patients | 0.240<sup>†</sup> | 0.037 | 76 | [31] |
|  | higher glomerular expression in nephrotic vs subnephrotic nephrosclerosis patients | 1.119* | 0.022 | 5 | [37] |
|  | negative correlation of tubulointerstitial expression with baseline proteinuria in MCD patients | -0.971<sup>†</sup> | 0.029 | 4 | [38] |
|  | lower glomerular expression in FSGS patients with nephrotic and subnephrotic proteinuria vs normal proteinuria | -1.435* | 0.047 | 91 | [31] |
|  | negative correlation of glomerular expression with baseline proteinuria in MCD patients | -0.222<sup>†</sup> | 0.047 | 80 | [31] |
|  | negative correlation of tubulointerstitial expression with blood pressure in healthy controls | -0.999<sup>†</sup> | 0.034 | 3 | [39] |
|  | negative correlation of glomerular expression with blood pressure in FSGS patients | -0.465<sup>†</sup> | 0.039 | 20 | [35] |
|  | lower glomerular expression in patients with *APOL1* high-risk genotype vs low-risk patients | -1.175* | 0.036 | 38 | [32] |
|  | negative correlation of glomerular expression with serum creatinine in healthy controls | -0.967<sup>†</sup> | 0.033 | 4 | [35] |
| C2CD4D | negative correlation of tubulointerstitial expression with eGFR in LN patients | -0.948<sup>†</sup> | 0.001 | 7 | ERCB |
|  | positive correlation of glomerular expression with baseline proteinuria in LN patients | 0.802<sup>†</sup> | 0.017 | 8 | ERCB |

\*log<sub>2</sub>(fold change); <sup>†</sup>r value/correlation coefficient; BUN, blood urea nitrogen; CKD, chronic kidney disease; DKD, diabetic kidney disease; eGFR, estimated glomerular filtration rate; ERCB, European Renal cDNA Bank; FSGS, focal segmental glomerulosclerosis; IgAN, IgA nephropathy; LN, lupus nephritis; MCD, minimal change disease

### Genetic association evidence for pathway-orphan genes in CKD

The GWAS Catalog was systematically queried for associations with kidney-relevant phenotypes and the results are summarized in **Table 3**. These genetic associations are based on predominantly European ancestry cohorts, such as the UK Biobank. Lists of cis-eQTLs within 1mbp of the pathway-orphan genes are provided in **Supplementary Table 7**. Using the significance threshold of *P*=5E-8, we have found four cis-eQTLs for *SEC14L6* in the glomerular tissue and 27 SNPs in the tubulointerstitial compartment based on NephQTL2 data.

**Table 3.**
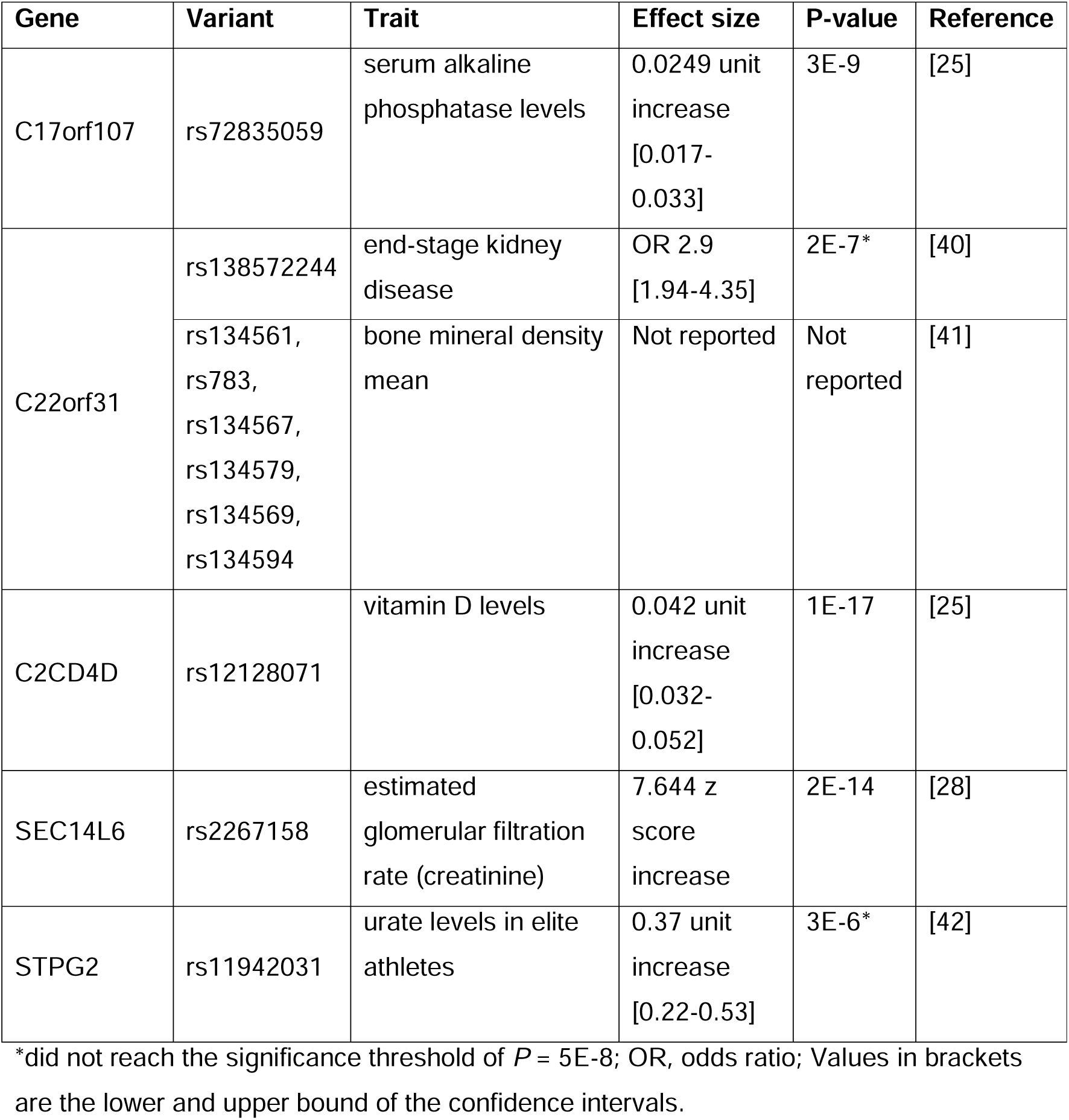
Variants mapping to pathway-orphan genes and their associated kidney-relevant traits.

| Gene | Variant | Trait | Effect size | P-value | Reference |
| --- | --- | --- | --- | --- | --- |
| C17orf107 | rs72835059 | serum alkaline phosphatase levels | 0.0249 unit increase<br>[0.017-0.033] | 3E-9 | [25] |
| C22orf31 | rs138572244 | end-stage kidney disease | OR 2.9<br>[1.94-4.35] | 2E-7* | [40] |
|  | rs134561, rs783, rs134567, rs134579, rs134569, rs134594 | bone mineral density mean | Not reported | Not reported | [41] |
| C2CD4D | rs12128071 | vitamin D levels | 0.042 unit increase<br>[0.032-0.052] | 1E-17 | [25] |
| SEC14L6 | rs2267158 | estimated glomerular filtration rate (creatinine) | 7.644 z score increase | 2E-14 | [28] |
| STPG2 | rs11942031 | urate levels in elite athletes | 0.37 unit increase<br>[0.22-0.53] | 3E-6* | [42] |

\*did not reach the significance threshold of $P = 5E-8$ ; OR, odds ratio; Values in brackets are the lower and upper bound of the confidence intervals.

## DISCUSSION

Pathway or gene set enrichment analysis has long become a routine step in the ‘-omics’ data bioinformatics analysis pipeline to infer the biological significance of genes or proteins found to be dysregulated in disease [1]. However, pathway analysis relies on predefined pathway annotations found in existing databases, thus making the method dependent on previously established pathway memberships. Moreover, some genes have been allocated to multiple pathways (e.g., complement factor C2 is included in 1 466 pathways and gene sets from ConsensusPathDB) resulting in their overrepresentation and inflated statistics, while some others are completely lacking pathway membership (‘pathway-orphan’ genes) and thus are routinely filtered out from the subsequent analysis. This study aimed to shed light on these previously overlooked genes in the kidney and their potential as an untapped source of novel disease biological insights.

We performed a data-driven exploration of the functionally enigmatic protein-coding genome focusing on those expressed in healthy kidneys and dysregulated in CKD, and observed that despite being uncharacterized, pathway-orphan genes have potential physiological roles in the kidney and pathophysiological function in CKD. Signals of kidney relevance include enriched expression in the kidney, significant modulation in CKD, and genetic associations with kidney function. These support our hypothesis that pathway-orphan genes warrant further functional characterization and mechanistic studies to determine how they can be involved in disease pathogenesis.

We report that many pathway-orphan genes are located in the X chromosome. Due to its high-sequence identity regions and transmission pattern that causes technical artifacts, the X chromosome is typically excluded from genomic analyses [43]. However, the X chromosome comprises 5% of the human genome and it also bears several genes involved in the immune response [43]. Moreover, the X chromosome is an important factor in disease, often contributing in a sex-specific manner. Pathway-orphan genes in the X chromosome thus deserve further studies, such as *MED14OS* as it is also dysregulated in CKD.

While pathway-orphan proteins are generally shorter and lighter than housekeeping and pathway-annotated proteins, they should not be overlooked as recent studies have shown how microproteins—proteins typically less than 100 amino acids in length—are involved in diverse biological processes. These microproteins encoded by short open reading frames can function as allosteric regulators of other proteins, signaling molecules, or effector proteins [44]. Eleven kidney-expressed pathway-orphan proteins are less than 100 amino acids long, which includes *SMIM35*. Other small integral membrane proteins (e.g. *SMIM22*, *SMIM43*) have been found to have roles as regulators of cytoskeletal organization and glucose transport [44].

Out of the 10 pathway-orphan genes dysregulated in CKD, *C10orf105* and *SEC14L6* have multiple lines of evidence for potential involvement in CKD. Both are upregulated at the mRNA level in diseased kidneys and are genetically linked to glomerular filtration rate, either through its regulatory enhancers (*C10orf105*) or its own genetic variants (*SEC14L6*). While genome-wide significant associations of gene variants to CKD or other kidney function traits do not confirm causation of pathology, they shed light on genes that might be involved in pathogenesis despite lack of mechanistic understanding. Single-pass membrane proteins are transmembrane proteins that play key roles in signal transduction, cell communication, immunity, transport, and energy conversion [45]. *C10orf105* has a transmembrane helix domain and is a single-pass membrane protein, which suggests its potential function. For *SEC14L6*, the presence of the CRAL-TRIO lipid binding domain could indicate a role in lipid traffic [26]. *SEC14L6* also has significant cis-eQTLs from glomeruli and tubulointerstitium derived from kidney biopsies of nephrotic syndrome patients.

Looking at which protein family these pathway-orphan genes proteins belong to and which protein domains they contain also gives us clues on their possible roles. The C2 domain in *C2CD4D*, in general, senses the cellular lipid microenvironment and can regulate lipid signal transduction and membrane trafficking, which could be its role in the kidney [46]. *IQANK1* has ankyrin repeats, commonly found in Notch receptors, which is involved in CKD pathophysiology [47]. There are seven pathway-orphan genes with experimentally validated interactors and one (*SEC14L6*) with a known ubiquitination site, suggesting that these proteins are part of protein networks and could serve a function.

Majority of the pathway-orphan genes are also expressed in the human fetal kidney (varying expression levels from Week 9 to 21) [12, 16], hinting about their involvement in kidney development. Defective kidney development has been linked to kidney disease and multiple evidence suggest that nephron deficiencies from possibly developmental defects can lead to CKD [48].

The remaining 24 pathway-orphan genes dysregulated in CKD could still have potential pathophysiological roles and are also worth investigating. Increasing the number of CKD RNA-seq studies analyzed could reveal additional evidence for these pathway-orphan genes.

We acknowledge several limitations of this study. Databases and annotation repositories are dynamic and are updated periodically, therefore, the validity of the results is restricted to the accession dates. Database identifiers can also be inaccurate, redundant, or obsolete. Some missingness accounts for a small proportion of protein-coding genes without expression level and functional data resulting in them having been missed in our analyses. Lastly, RNA-seq-based gene expression detection is dependent upon the sequencing depth, increasing this might potentially reveal additional low-abundant transcripts.

Despite these limitations, our data-driven investigation of this overlooked group of genes uses an analytical approach to characterize them. Previous studies have reiterated that data-driven hypotheses and analyses can help alleviate the annotation inequality we observe in all these databases [1, 3]. Our design should also apply to other tissues and organs, as well as other diseases.

In conclusion, a number of routinely overlooked ‘pathway-orphan’ genes are likely to have a plausible link to kidney physiology or play a role in disease. We hope that our results will help rectify a bias in functional gene annotation and spark subsequent hypothesis-testing investigations, for example, through mechanistic experimental or Mendelian Randomization type of studies, that are warranted to verify their potential roles in CKD pathogenesis.

## Data availability statement

All data analyzed in this study are publicly available and cited within the manuscript.

## Acknowledgements

Initial preliminary results from this work were previously shared as an abstract at the ASN Kidney Week 2022 (November 3 – 6, 2022) and as an abstract and poster at the World Congress of Nephrology 2023 (March 30 – April 2, 2023).

## Funding

This research was supported by the European Union Horizon 2020 research and innovation programme under the Marie Skłodowska-Curie grant agreement No. 860977 titled TrainCKDis.

## Authors‘ contributions

Dianne Acoba: Data curation, Formal analysis, Investigation, Methodology, Resources, Software, Validation, Visualization, Writing – original draft, Writing – review & editing

Sophie Gunnarsson: Data curation, Formal analysis, Investigation, Methodology, Resources, Software, Visualization, Writing – review & editing

Anna Reznichenko: Conceptualization, Funding acquisition, Project administration, Resources, Supervision, Writing – review & editing

## Conflict of interest statement

DA, SG, and AR are AstraZeneca employees. DA is an industrial PhD student at AstraZeneca.

## Notes

### Summary of Updates

Abstract was updated for easier reading.

