## Supplementary Materials for "Kidney ‘pathway-orphan’ genes as a potential overlooked source of novel insights into kidney health and disease"

**Supplementary Table 1.** Dataset versions utilized in the study.

**Supplementary Table 2.** Information on human protein-coding genes. (Provided as a separate supplemental spreadsheet.)

**Supplementary Table 3.** Count of protein-coding genes in each subcellular compartment as predicted by DeepLoc.

**Supplementary Table 4.** Protein domains in pathway-orphan genes as predicted by Pfam through InterPro.

**Supplementary Table 5.** The 97 kidney-expressed pathway-orphan genes.

**Supplementary Table 6.** List of variants mapping to within 1 megabase pairs of the pathway-orphan genes from NephQTL2.

**Supplementary References**

**Supplementary Table 1. Dataset versions utilized in the study.**

| Dataset | Source | Date Updated |
|---|---|---|
| Protein-coding genes | HGNC [1] | 26 January 2024 |
| Gene Ontology (Biological Process and Molecular Function) | Gene Ontology [2, 3] | 17 January 2024 |
| Reactome v87 | Reactome [4] | 4 December 2023 |
| KEGG v109 | KEGG [5] | 1 January 2024 |
| Hallmark and Curated Gene Sets | MSigDB [6, 7] | October 2023 |
| WikiPathways | WikiPathways [8] | 11 January 2024 |
| Ingenuity Pathway Analysis | QIAGEN [9] | December 2023 |
| CPDB v35 | ConsensusPathDB [10] | 5 June 2021 |
| GENCODE | GENCODE [11] | January 2024 |
| UniProt | UniProt [12] | 24 January 2024 |
| InterPro v98.0 | InterPro [13] | 25 January 2024 |
| DeepLoc 2.026 | DeepLoc [14] | 2022 |
| gene2pubmed | NCBI | 26 January 2024 |
| GTEx v8 | GTEx [15] | 5 June 2017 |
| HPA v23.0 | HPA [16] | 19 June 2023 |
| DESCARTES | DESCARTES [17] | 13 November 2020 |
| Bgee v15.1 | Bgee [18] | 11 November 2023 |
| GWAS Catalog | GWAS Catalog [19] | 19 January 2024 |
| HumanBase | HumanBase [20] | July 2022 |
| BioGrid 4.4.229 | BioGrid [21] | January 2024 |
| AGR 6.0 | AGR [22] | 29 September 2023 |
| Pharos 3.18.0 | Pharos [23] | 12 October 2023 |
| Nephroseq v5 | Nephroseq [24] | March 2023 |
| GeneHancer 5.19 | GeneCards [25] | 15 January 2024 |
| NephQTL2 | NephQTL2 [26] | 2023 |

**Supplementary Table 3. Count of protein-coding genes in each subcellular compartment as predicted by DeepLoc.**

| Localizations | Pathway-orphan genes (%) | Housekeeping genes (%) | Pathway-annotated genes (%) |
|---|---|---|---|
| Cytoplasm | 68 (24%) | 110 (27%) | 4081 (22%) |
| Nucleus | 63 (22%) | 113 (28%) | 3116 (17%) |
| Cytoplasm\|Nucleus | 36 (13%) | 180 (44%) | 2564 (14%) |
| Cell membrane | 29 (10%) | 0 (0%) | 2740 (15%) |
| Endoplasmic reticulum | 19 (7%) | 2 (0.5%) | 908 (5%) |
| Extracellular | 19 (7%) | 0 (0%) | 1451 (8%) |
| Cell membrane\|Lysosome/Vacuole | 4 (1%) | 0 (0%) | 548 (3%) |
| Lysosome/Vacuole | 4 (1%) | 0 (0%) | 116 (0.6%) |
| Mitochondrion | 4 (1%) | 1 (0.2%) | 935 (5%) |
| Cytoplasm\|Cell membrane | 3 (1%) | 0 (0%) | 493 (3%) |
| Cytoplasm\|Endoplasmic reticulum | 2 (0.7%) | 0 (0%) | 27 (0.1%) |
| Endoplasmic reticulum\|Lysosome/Vacuole | 2 (0.7%) | 0 (0%) | 45 (0.2%) |
| Nucleus\|Cell membrane | 2 (0.7%) | 0 (0%) | 1 (0.005%) |
| Cell membrane\|Endoplasmic reticulum\|Lysosome/Vacuole | 1 (0.3%) | 0 (0%) | 31 (0.2%) |
| Cytoplasm\|Extracellular | 1 (0.3%) | 0 (0%) | 29 (0.2%) |
| Endoplasmic reticulum\|Golgi apparatus | 1 (0.3%) | 0 (0%) | 125 (0.7%) |
| Extracellular\|Cell membrane\|Lysosome/Vacuole | 1 (0.3%) | 0 (0%) | 1 (0.005%) |
| Peroxisome | 1 (0.3%) | 0 (0%) | 44 (0.2%) |
| Cytoplasm\|Lysosome/Vacuole | 0 (0%) | 1 (0.2%) | 237 (1%) |

**Supplementary Table 4. Protein domains in pathway-orphan genes as predicted by Pfam through InterPro.**

| Protein domain | Gene count | Genes |
|---|---|---|
| Unknown function | 28 | C1orf141, C1orf167, C1orf185, C2orf78, C4orf51, C5orf52, C8orf89, C9orf57, C10orf105, C12orf71, C16orf82, C17orf107, C18orf63, C22orf31, CXorf49B, CCDC177, KIAA2012, NBPF6, NBPF19, NBPF26, PROB1, PRR23A, PRR23C, PRR23D2, PRR33, TEX46, TEX52, TMEM217B |
| Zinc knuckle | 18 | FAM90A3, FAM90A5, FAM90A7, FAM90A8, FAM90A11, FAM90A12, FAM90A13, FAM90A14, FAM90A15, FAM90A16, FAM90A17, FAM90A18, FAM90A19, FAM90A20, FAM90A22, FAM90A23, FAM90A24, FAM90A26 |
| Nuclear pore complex interacting protein (NPIP) | 9 | NPIPA2, NPIPA3, NPIPA5, NPIPA8, NPIPA9, NPIPB2, NPIPB6, NPIPB8, NPIPB9 |
| GAGE protein | 7/9 | GAGE2C, GAGE6, GAGE8, GAGE12C (GAGE12D, GAGE12E), GAGE12H, GAGE12J, PAGE2B |
| INTS6/SAGE1/DDX26B/CT45 C-terminus | 6 | CT45A2, CT45A5, CT45A6, CT45A7, CT45A8, CT45A9 |
| Ankyrin repeat | 5 | ANKRD30BL, ANKRD60, ANKRD62, IQANK1, POTEC |
| Cancer/testis gene family 47 | 3 | CT47A1, CT47B1, CT47C1 |
| WD domain, G-beta repeat | 3 | DCAF8L1, DCAF8L2, WDR88 |
| B melanoma antigen family | 2 | BAGE3, BAGE4 |
| ENV polyprotein (coat polyprotein) | 2 | ERVV-1, ERVV-2 |
| FAM194 protein | 2 | C3orf20, ERICH6B |
| Interferon-induced transmembrane protein | 2 | PMIS2, PRRT1B |
| Leucine rich repeat | 2 | LRRC3C, LRRC53 |

| | | |
|---|---|---|
| NUT protein | 2 | NUTM2B, NUTM2G |
| PNMA | 2 | PNMA6E, PNMA6F |
| Proline-rich protein family 20 | 2 | PRR20B, PRR20G |
| Sperm protein associated with nucleus, mapped to X chromosome | 2 | SPANXN4, SPANXN5 |
| u-PAR/Ly-6 domain | 2 | LY6L, LY6S |
| ARF7 effector protein C-terminus | 1 | ARL14EPL |
| Armadillo-like helical domain-containing protein 2 | 1 | ARMH2 |
| Beta-lactamase | 1 | LACTBL1 |
| Borna disease virus P40 protein | 1 | EBLN1 |
| C-type lysozyme/alpha-lactalbumin family | 1 | SPACA5B |
| C2 domain | 1 | C2CD4D |
| CCDC144C protein coiled-coil region | 1 | POTEB2 |
| CRAL/TRIO domain | 1 | SEC14L6 |
| CUB domain | 1 | SPADH |
| Cardiac transcription factor regulator, Developmental protein | 1 | LBHD2 |
| Cation channel sperm-associated auxiliary subunit TMEM249 | 1 | TMEM249 |
| Chibby family | 1 | CBY3 |
| Cortexin of kidney | 1 | CTXND2 |
| Cyclin, N-terminal domain | 1 | CCNYL1B |
| Doublesex-and mab-3-related transcription factor C1 and C2 | 1 | DMRTC1B |
| Exocyst complex component SEC3 N-terminal PIP2 binding PH | 1 | EXOC1L |
| FAM153 family | 1 | FAM153B |
| FAM24 family | 1 | FAM24A |
| FAM25 family | 1 | FAM25G |

| | | |
|---|---|---|
| Facioscapulohumeral muscular dystrophy candidate 2 | 1 | FRG2C |
| Fibronectin type III domain-containing protein 10 | 1 | FNDC10 |
| GM130 C-terminal binding motif | 1 | GOLGA6L7 |
| GSG1-like protein | 1 | GSG1L2 |
| HSF-type DNA-binding | 1 | HSFY2 |
| Hemingway/CFA97 | 1 | CFAP97D2 |
| Immunoglobulin I-set domain | 1 | SPEGNB |
| Immunoglobulin V-set domain | 1 | FAM187A |
| Keratin, high sulfur B2 protein | 1 | KRTAP4-16 |
| Kinocilin protein | 1 | KNCN |
| Kita-kyushu lung cancer antigen 1 | 1 | CT83 |
| Low-density lipoprotein receptor domain class A | 1 | LDLRAD2 |
| MORN repeat | 1 | RSPH10B2 |
| MyoD family inhibitor | 1 | MDFIC2 |
| PGC7/Stella/Dppa3 domain | 1 | FAM156B |
| PMP-22/EMP/MP20/Claudin tight junction | 1 | TMEM235 |
| PNMA N-terminal RRM-like domain | 1 | PNMA8C |
| Putative golgin subfamily A member 2-like protein 5 | 1 | GOLGA6L10 |
| Rab-GTPase-TBC domain | 1 | TBC1D28 |
| RanBP1 domain | 1 | RGPD6 |
| Reeler domain | 1 | REELD1 |
| SH2 domain | 1 | SH2D7 |
| Sperm-tail PG-rich repeat | 1 | STPG2 |
| TLR4 regulator and MIR-interacting MSAP | 1 | CNPY1 |
| TMCO5 family | 1 | TMEM191C |
| TMEM210 family | 1 | TMEM210 |
| TMEM240 family | 1 | TMEM240 |
| Tetraspanin family | 1 | TSPAN19 |

| | | |
|---|---|---|
| Translation machinery associated TMA7 | 1 | TMA7B |
| Transmembrane protein 247 | 1 | TMEM247 |
| UPF0731 family | 1 | FAM229A |
| Variable charge X/Y family | 1 | VCY1B |
| zinc finger of C3HC4-type, RING | 1 | TRIM49D2 |

**Supplementary Table 5. The 97 kidney-expressed pathway-orphan genes.**

| Ensembl gene ID | Symbol | Gene name | Chromosome | Gene length | TPM Cortex | TPM Medulla | Z-score Cortex | Z-score Medulla | Protein length | Protein mass | Publication Count |
|---|---|---|---|---|---|---|---|---|---|---|---|
| ENSG00000215910 | C1orf167 | chromosome 1 open reading frame 167 | 1 | 27798 | 0.01 | 0.02 | 0.29 | -0.10 | 1468 | 162423 | 13 |
| ENSG00000225556 | C2CD4D | C2 calcium dependent domain containing 4D | 1 | 2739 | 0.63 | 0.64 | 0.33 | -0.07 | 353 | 37583 | 3 |
| ENSG00000131379 | C3orf20 | chromosome 3 open reading frame 20 | 3 | 97895 | 0.04 | 0.07 | 0.28 | -0.10 | 904 | 101266 | 14 |
| ENSG00000180697 | C3orf22 | chromosome 3 open reading frame 22 | 3 | 31966 | 0.02 | 0.04 | 0.30 | -0.10 | 141 | 15686 | 4 |
| ENSG00000163632 | C3orf49 | chromosome 3 open reading frame 49 | 3 | 29337 | 0.05 | 0.07 | -0.24 | -0.41 | 292 | 33461 | 6 |
| ENSG00000237136 | C4orf51 | chromosome 4 open reading frame 51 | 4 | 90886 | 0.05 | 0.05 | 0.33 | -0.07 | 202 | 23001 | 1 |
| ENSG00000234511 | C5orf58 | chromosome 5 open reading frame 58 | 5 | 23471 | 0.15 | 0.14 | 0.24 | -0.14 | 81 | 9231 | 3 |
| ENSG00000204661 | C5orf60 | chromosome 5 open reading frame 60 | 5 | 3502 | 0.05 | 0.06 | 0.30 | -0.10 | 353 | 39250 | 7 |
| ENSG00000274443 | C8orf89 | chromosome 8 open reading frame 89 | 8 | 18199 | 0.13 | 0.25 | 0.18 | -0.12 | 161 | 18160 | 1 |

| | | | | | | | | | | | |
|---|---|---|---|---|---|---|---|---|---|---|---|
| ENSG00000214688 | C10orf105 | chromosome 10 open reading frame 105 | 10 | 26123 | 0.04 | 0.09 | -0.30 | -0.49 | 133 | 14519 | 2 |
| ENSG00000204365 | C10orf126 | chromosome 10 open reading frame 126 | 10 | NA | 0.45 | 0.32 | 4.47 | 1.94 | 172 | 19474 | 3 |
| ENSG00000214700 | C12orf71 | chromosome 12 open reading frame 71 | 12 | 1521 | 0.10 | 0.16 | 0.02 | -0.22 | 269 | 30355 | 1 |
| ENSG00000226792 | C13orf42 | chromosome 13 open reading frame 42 | 13 | 118133 | 0.01 | 0.12 | 0.17 | 0.30 | 325 | 37385 | 1 |
| ENSG00000283199 | C13orf46 | chromosome 13 open reading frame 46 | 13 | 20371 | 0.12 | 0.11 | -0.01 | -0.31 | 212 | 23433 | 1 |
| ENSG00000205710 | C17orf107 | chromosome 17 open reading frame 107 | 17 | 3516 | 1.13 | 1.54 | -0.04 | -0.21 | 190 | 19931 | 0 |
| ENSG00000267221 | C17orf113 | chromosome 17 open reading frame 113 | 17 | 12369 | 1.94 | 2.97 | 0.12 | 0.37 | 675 | 72432 | 0 |
| ENSG00000262165 | C17orf114 | chromosome 17 open reading frame 114 | 17 | 5854 | 0.49 | 0.25 | -0.73 | -1.06 | 79 | 8292 | 0 |
| ENSG00000100249 | C22orf31 | chromosome 22 open reading frame 31 | 22 | 3159 | 0.20 | 0.25 | 0.20 | -0.15 | 290 | 32655 | 12 |
| ENSG00000204659 | CBY3 | chibby family member 3 | 5 | 2474 | 0.25 | 0.35 | 0.09 | -0.21 | 242 | 27343 | 2 |
| ENSG00000230561 | CCDC192 | coiled-coil domain containing 192 | 5 | 238146 | 0.15 | 0.21 | 0.11 | -0.20 | 292 | 32479 | 4 |
| ENSG00000269720 | CCDC194 | coiled-coil domain containing 194 | 19 | 7000 | 0.22 | 0.20 | 0.21 | -0.17 | 234 | 24954 | 0 |

| | | | | | | | | | | | |
|---|---|---|---|---|---|---|---|---|---|---|---|
| ENSG00000236383 | CCDC200 | coiled-coil domain containing 200 | 17 | 88456 | 1.48 | 1.03 | 0.47 | -0.07 | 168 | 19413 | 3 |
| ENSG00000223510 | CDRT15 | CMT1A duplicated region transcript 15 | 17 | 1189 | 0.07 | 0.24 | 0.27 | -0.05 | 188 | 20651 | 6 |
| ENSG00000283361 | CFAP97D2 | CFAP97 domain containing 2 | 13 | 68394 | 0.02 | 0.06 | -0.16 | -0.35 | 98 | 11681 | 1 |
| ENSG00000283324 | CTXND2 | cortexin domain containing 2 | 1 | 26156 | 0.00 | 0.04 | 0.24 | 0.22 | 55 | 5988 | 2 |
| ENSG00000184911 | DMRTC1B | DMRT like family C1B | X | 71913 | 0.02 | 0.04 | 0.01 | -0.14 | 192 | 20139 | 6 |
| ENSG00000165837 | ERICH6B | glutamate rich 6B | 13 | 81217 | 0.12 | 0.20 | -0.01 | -0.20 | 696 | 81686 | 8 |
| ENSG00000269526 | ERVV-1 | endogenous retrovirus group V member 1, envelope | 19 | 2202 | 0.00 | 0.55 | 0.04 | 1.71 | 477 | 52557 | 9 |
| ENSG00000268964 | ERVV-2 | endogenous retrovirus group V member 2, envelope | 19 | 6940 | 0.00 | 0.32 | 0.23 | 7.33 | 535 | 59317 | 9 |
| ENSG00000277150 | F8A3 | coagulation factor VIII associated 3 | X | 1706 | 0.03 | 0.01 | -0.84 | -2.08 | 371 | 39103 | 8 |
| ENSG00000188859 | FAM78B | family with sequence similarity 78 member B | 1 | 109575 | 0.58 | 0.62 | -0.58 | -0.68 | 261 | 29835 | 10 |
| ENSG00000233295 | FAM90A20 | family with sequence similarity 90 member A20 | 8 | 3010 | 0.06 | 0.08 | -0.96 | -0.81 | 464 | 50026 | 3 |

| | | | | | | | | | | | |
|---|---|---|---|---|---|---|---|---|---|---|---|
| ENSG00000182230 | FAM153B | family with sequence similarity 153 member B | 5 | NA | 1.35 | 0.97 | -0.08 | -0.40 | 387 | 43591 | 8 |
| ENSG00000179304 | FAM156B | family with sequence similarity 156 member B | X | 17254 | 0.03 | 0.03 | -0.32 | -0.87 | 213 | 24412 | 8 |
| ENSG00000225828 | FAM229A | family with sequence similarity 229 member A | 1 | 3008 | 11.28 | 11.69 | 0.29 | -0.10 | 127 | 12972 | 4 |
| ENSG00000283267 | FAM237B | family with sequence similarity 237 member B | 7 | 4801 | 0.19 | 0.37 | 0.43 | 0.99 | 139 | 16331 | 2 |
| ENSG00000283473 | FAM240A | family with sequence similarity 240 member A | 3 | 14018 | 0.17 | 0.19 | 3.68 | 2.52 | 83 | 10377 | 1 |
| ENSG00000216921 | FAM240C | family with sequence similarity 240 member C | 2 | 8563 | 0.07 | 0.19 | -0.63 | -0.68 | 95 | 10834 | 2 |
| ENSG00000228594 | FNDC10 | fibronectin type III domain containing 10 | 1 | 2123 | 3.49 | 3.20 | -0.29 | -0.54 | 226 | 24218 | 4 |
| ENSG00000206262 | FOXL2NB | FOXL2 neighbor | 3 | 6773 | 0.00 | 0.02 | 0.08 | -0.24 | 175 | 18625 | 2 |
| ENSG00000172969 | FRG2C | FSHD region gene 2 family member C | 3 | 2892 | 0.04 | 1.39 | 0.31 | 4.26 | 282 | 30798 | 3 |
| ENSG00000278662 | GOLGA6L10 | golgin A6 family like 10 | 15 | 9482 | 0.33 | 0.71 | -0.23 | -0.21 | 522 | 60902 | 4 |

| | | | | | | | | | | | |
|---|---|---|---|---|---|---|---|---|---|---|---|
| ENSG00000228789 | HCG22 | HLA complex group 22 (gene/pseudogene) | 6 | 6440 | 0.03 | 0.09 | 0.14 | -0.20 | 251 | 26282 | 14 |
| ENSG00000203499 | IQANK1 | IQ motif and ankyrin repeat containing 1 | 8 | 56506 | 1.20 | 2.69 | 0.09 | -0.04 | 560 | 63562 | 20 |
| ENSG00000182329 | KIAA2012 | KIAA2012 | 2 | 131933 | 0.01 | 0.03 | -0.24 | -0.43 | 1180 | 135305 | 5 |
| ENSG00000283071 | LBHD2 | LBH domain containing 2 | 14 | 5817 | 0.10 | 0.17 | -0.08 | -0.30 | 108 | 10701 | 0 |
| ENSG00000187942 | LDLRAD2 | low density lipoprotein receptor class A domain containing 2 | 1 | 12960 | 1.03 | 1.17 | -0.46 | -0.58 | 272 | 28581 | 3 |
| ENSG00000261667 | LY6L | lymphocyte antigen 6 family member L | 8 | 2856 | 0.85 | 0.14 | 10.51 | 1.00 | 138 | 15317 | 1 |
| ENSG00000234636 | MED14OS | MED14 opposite strand | X | 3301 | 1.72 | 2.67 | 1.31 | 2.15 | 135 | 14289 | 1 |
| ENSG00000283683 | MYOCOS | myocilin opposite strand | 1 | 38178 | 0.13 | 0.16 | 4.06 | 3.20 | 80 | 8950 | 0 |
| ENSG00000271383 | NBPF19 | NBPF member 19 | 1 | 81316 | 0.60 | 1.22 | -1.16 | -0.65 | 3843 | 440408 | 8 |
| ENSG00000273136 | NBPF26 | NBPF member 26 | 1 | 118284 | 0.86 | 1.81 | -1.17 | -0.65 | 902 | 103816 | 11 |
| ENSG00000254852 | NPIPA2 | nuclear pore complex interacting protein family member A2 | 16 | 22929 | 0.66 | 1.06 | -0.46 | -0.34 | 369 | 42223 | 3 |
| ENSG00000224712 | NPIPA3 | nuclear pore complex interacting | 16 | 22931 | 0.08 | 0.20 | -0.58 | -0.07 | 350 | 40061 | 3 |

| | | | | | | | | | | | |
|---|---|---|---|---|---|---|---|---|---|---|---|
| | | protein family member A3 | | | | | | | | | |
| ENSG00000183793 | NPIPA5 | nuclear pore complex interacting protein family member A5 | 16 | 17419 | 3.89 | 8.40 | -0.41 | -0.18 | 350 | 40105 | 4 |
| ENSG00000183889 | NPIPA6 | nuclear pore complex interacting protein family, member A6 | 16 | 33146 | 1.23 | 1.10 | -0.68 | -0.82 | NA | NA | 1 |
| ENSG00000214940 | NPIPA8 | nuclear pore complex interacting protein family member A8 | 16 | 18817 | 0.03 | 0.05 | -0.17 | -0.25 | 369 | 42230 | 3 |
| ENSG00000233024 | NPIPA9 | nuclear pore complex interacting protein family, member A9 | 16 | 21245 | 0.14 | 0.23 | -0.36 | -0.29 | 369 | 42104 | 2 |
| ENSG00000234719 | NPIPB2 | nuclear pore complex interacting protein family member B2 | 16 | 49384 | 1.19 | 2.52 | 0.39 | 0.62 | 397 | 45575 | 3 |
| ENSG00000198156 | NPIPB6 | nuclear pore complex interacting protein family member B6 | 16 | 20991 | 0.25 | 0.27 | 0.05 | -0.26 | 425 | 49162 | 5 |
| ENSG00000196993 | NPIPB9 | nuclear pore complex interacting | 16 | 21077 | 0.05 | 0.01 | -0.41 | -0.87 | 429 | 49181 | 4 |

| | | | | | | | | | | | |
|---|---|---|---|---|---|---|---|---|---|---|---|
| | | protein family member B9 | | | | | | | | | |
| ENSG00000188199 | NUTM2B | NUT family member 2B | 10 | 11454 | 0.09 | 0.27 | -0.35 | 0.96 | 878 | 93984 | 5 |
| ENSG00000188152 | NUTM2G | NUT family member 2G | 9 | 11943 | 0.08 | 0.15 | 0.09 | -0.05 | 741 | 79011 | 2 |
| ENSG00000238269 | PAGE2B | PAGE family member 2B | X | 3879 | 0.11 | 0.15 | 0.28 | -0.11 | 111 | 12041 | 5 |
| ENSG00000277531 | PNMA8C | PNMA family member 8C | 19 | 4239 | 0.03 | 0.15 | -0.32 | -0.51 | 204 | 22873 | 0 |
| ENSG00000228672 | PROB1 | proline rich basic protein 1 | 5 | 4512 | 1.63 | 1.49 | 0.02 | -0.31 | 1015 | 106917 | 3 |
| ENSG00000283787 | PRR33 | proline rich 33 | 11 | 3817 | 0.42 | 0.54 | -0.34 | -0.53 | 331 | 35187 | 4 |
| ENSG00000283526 | PRRT1B | proline rich transmembrane protein 1B | 9 | 14292 | 0.29 | 0.71 | 0.53 | 0.68 | 263 | 26769 | 1 |
| ENSG00000224940 | PRRT4 | proline rich transmembrane protein 4 | 7 | 11365 | 0.06 | 0.06 | -0.64 | -0.74 | 899 | 92712 | 10 |
| ENSG00000250673 | REELD1 | reeler domain containing 1 | 4 | 17752 | 0.73 | 1.59 | 0.06 | 0.84 | 526 | 56837 | 3 |
| ENSG00000169402 | RSPH10B2 | radial spoke head 10 homolog B2 | 7 | 45262 | 0.01 | 0.02 | 0.25 | -0.10 | 870 | 100547 | 7 |
| ENSG00000268655 | SAXO3 | stabilizer of axonemal microtubules 3 | 19 | 2581 | 0.26 | 0.36 | 0.31 | 0.01 | 334 | 37135 | 0 |
| ENSG00000214491 | SEC14L6 | SEC14 like lipid binding 6 | 22 | 23942 | 6.60 | 7.67 | 3.45 | 2.55 | 397 | 45364 | 3 |
| ENSG00000183476 | SH2D7 | SH2 domain containing 7 | 15 | 26562 | 0.06 | 0.08 | 0.09 | -0.22 | 451 | 49807 | 3 |
| ENSG00000165935 | SMCO2 | single-pass membrane protein with | 12 | 55449 | 0.08 | 0.23 | -0.46 | -0.26 | 343 | 39487 | 2 |

| | | | | | | | | | | | |
|---|---|---|---|---|---|---|---|---|---|---|---|
| | | coiled-coil domains 2 | | | | | | | | | |
| ENSG00000255274 | SMIM35 | small integral membrane protein 35 | 11 | 84665 | 0.18 | 0.09 | 1.05 | -0.16 | 85 | 9379 | 0 |
| ENSG00000221843 | SPATA31H1 | SPATA31 subfamily H member 1 | 2 | 45336 | 0.12 | 0.28 | 0.20 | -0.12 | 1984 | 224321 | 15 |
| ENSG00000185523 | SPATA45 | spermatogenesis associated 45 | 1 | 17508 | 0.14 | 0.22 | 0.26 | -0.10 | 98 | 11356 | 2 |
| ENSG00000226763 | SRRM5 | serine/arginine repetitive matrix 5 | 19 | 17881 | 1.09 | 2.09 | 0.27 | -0.05 | 715 | 80355 | 4 |
| ENSG00000163116 | STPG2 | sperm tail PG-rich repeat containing 2 | 4 | 959383 | 0.14 | 0.21 | 1.38 | 1.40 | 459 | 50660 | 8 |
| ENSG00000197768 | STPG3 | sperm-tail PG-rich repeat containing 3 | 9 | 2222 | 3.04 | 2.71 | 0.49 | 0.01 | 386 | 42496 | 7 |
| ENSG00000189375 | TBC1D28 | TBC1 domain family member 28 | 17 | 10180 | 0.00 | 0.03 | 0.29 | -0.10 | 210 | 24072 | 6 |
| ENSG00000227868 | TEX46 | testis expressed 46 | 1 | 8266 | 0.08 | 0.17 | 0.29 | -0.10 | 121 | 14053 | 0 |
| ENSG00000283297 | TEX52 | testis expressed 52 | 12 | 7993 | 0.51 | 0.63 | 0.08 | -0.21 | 305 | 35334 | 0 |
| ENSG00000283268 | TEX54 | testis expressed 54 | 11 | 520 | 0.22 | 0.70 | 0.29 | -0.06 | 124 | 14093 | 0 |
| ENSG00000225528 | TMA7B | translation machinery associated 7 homolog B | 22 | 4508 | 0.07 | 0.09 | 0.08 | -0.20 | 64 | 7091 | 0 |
| ENSG00000177800 | TMEM78 | transmembrane protein 78 | 1 | 2174 | 0.00 | 0.03 | -0.50 | 0.43 | 136 | 15193 | 3 |

| | | | | | | | | | | | |
|---|---|---|---|---|---|---|---|---|---|---|---|
| ENSG00000206140 | TMEM191C | transmembrane protein 191C | 22 | 4846 | 0.40 | 0.30 | 0.24 | -0.16 | 302 | 34809 | 2 |
| ENSG00000205090 | TMEM240 | transmembrane protein 240 | 1 | 5846 | 1.23 | 1.81 | -0.59 | -0.66 | 173 | 19908 | 13 |
| ENSG00000261587 | TMEM249 | transmembrane protein 249 | 8 | 2735 | 0.45 | 1.10 | 0.07 | -0.19 | 235 | 27046 | 1 |
| ENSG00000274386 | TMEM269 | transmembrane protein 269 | 1 | 31628 | 0.08 | 0.10 | -0.29 | -0.45 | 203 | 22507 | 1 |
| ENSG00000281106 | TMEM272 | transmembrane protein 272 | 13 | 31830 | 0.08 | 0.10 | -0.02 | -0.31 | 187 | 21340 | 3 |
| ENSG00000205456 | TP53TG3D | TP53 target 3D | 16 | 2642 | 0.02 | 0.10 | 0.04 | -0.16 | 124 | 12828 | 3 |
| ENSG00000204025 | TRPC5OS | TRPC5 opposite strand | X | 27939 | 0.05 | 0.08 | 0.30 | -0.09 | 111 | 12326 | 6 |
| ENSG00000231738 | TSPAN19 | tetraspanin 19 | 12 | 21966 | 0.10 | 0.15 | -0.03 | -0.28 | 248 | 28460 | 4 |
| ENSG00000166359 | WDR88 | WD repeat domain 88 | 19 | 43709 | 0.24 | 0.48 | 0.20 | -0.07 | 472 | 52621 | 5 |
| ENSG00000236782 | ZNF593OS | ZNF593 opposite strand | 1 | 2316 | 0.16 | 0.20 | -0.70 | -0.67 | 63 | 6929 | 0 |

**Supplementary Table 6. List of variants mapping to within 1 megabase pairs of the pathway-orphan genes from NephQTL2.**

| rsID | Ref/Alt Allele | Alt Frequency | Beta | P-Value |
|---|---|---|---|---|
| ***SEC14L6*** **(Glomeruli)** | | | | |
| rs5749123 | G/A | 0.765 | 0.475 | 2.54E-10 |
| rs5749123 | G/A | 0.765 | 0.475 | 2.54E-10 |
| rs5749123 | G/A | 0.765 | 0.475 | 2.54E-10 |
| rs5753190 | T/C | 0.769 | 0.461 | 8.23E-10 |
| rs5753190 | T/C | 0.769 | 0.461 | 8.23E-10 |
| rs5753190 | T/C | 0.769 | 0.461 | 8.23E-10 |
| rs4820865 | A/G | 0.769 | 0.461 | 8.23E-10 |
| rs4820865 | A/G | 0.769 | 0.461 | 8.23E-10 |
| rs4820865 | A/G | 0.769 | 0.461 | 8.23E-10 |
| rs5753193 | C/G | 0.76 | 0.416 | 1.51E-08 |
| rs5753193 | C/G | 0.76 | 0.416 | 1.51E-08 |
| rs5753193 | C/G | 0.76 | 0.416 | 1.51E-08 |
| ***SEC14L6*** **(Tubules)** | | | | |
| rs4820865 | A/G | 0.759 | 0.74 | 1.42E-34 |
| rs4820865 | A/G | 0.759 | 0.74 | 1.42E-34 |
| rs4820865 | A/G | 0.759 | 0.74 | 1.42E-34 |
| rs5753190 | T/C | 0.762 | 0.739 | 1.35E-33 |
| rs5753190 | T/C | 0.762 | 0.739 | 1.35E-33 |
| rs5753190 | T/C | 0.762 | 0.739 | 1.35E-33 |
| rs5749123 | G/A | 0.757 | 0.741 | 7.34E-33 |
| rs5749123 | G/A | 0.757 | 0.741 | 7.34E-33 |
| rs5749123 | G/A | 0.757 | 0.741 | 7.34E-33 |
| rs5753193 | C/G | 0.749 | 0.709 | 3.30E-32 |
| rs5753193 | C/G | 0.749 | 0.709 | 3.30E-32 |
| rs5753193 | C/G | 0.749 | 0.709 | 3.30E-32 |
| rs5749120 | G/A | 0.737 | 0.649 | 2.42E-26 |
| rs5749120 | G/A | 0.737 | 0.649 | 2.42E-26 |
| rs5749120 | G/A | 0.737 | 0.649 | 2.42E-26 |
| rs2899151 | T/C | 0.704 | 0.585 | 1.72E-22 |
| rs2899151 | T/C | 0.704 | 0.585 | 1.72E-22 |
| rs2899151 | T/C | 0.704 | 0.585 | 1.72E-22 |

| | | | | |
|---|---|---|---|---|
| rs2079312 | A/G | 0.693 | 0.553 | 1.36E-19 |
| rs2079312 | A/G | 0.693 | 0.553 | 1.36E-19 |
| rs2079312 | A/G | 0.693 | 0.553 | 1.36E-19 |
| rs7285565 | A/G | 0.69 | 0.547 | 5.20E-19 |
| rs7285565 | A/G | 0.69 | 0.547 | 5.20E-19 |
| rs7285565 | A/G | 0.69 | 0.547 | 5.20E-19 |
| rs4820862 | A/G | 0.687 | 0.548 | 9.72E-19 |
| rs4820862 | A/G | 0.687 | 0.548 | 9.72E-19 |
| rs4820862 | A/G | 0.687 | 0.548 | 9.72E-19 |
| rs5994317 | G/A | 0.638 | 0.507 | 1.04E-18 |
| rs5994317 | G/A | 0.638 | 0.507 | 1.04E-18 |
| rs5994317 | G/A | 0.638 | 0.507 | 1.04E-18 |
| rs5753201 | C/G | 0.744 | 0.566 | 1.74E-18 |
| rs5753201 | C/G | 0.744 | 0.566 | 1.74E-18 |
| rs5753201 | C/G | 0.744 | 0.566 | 1.74E-18 |
| rs5753186 | A/G | 0.686 | 0.54 | 2.60E-18 |
| rs5753186 | A/G | 0.686 | 0.54 | 2.60E-18 |
| rs5753186 | A/G | 0.686 | 0.54 | 2.60E-18 |
| rs1107844 | T/C | 0.686 | 0.53 | 4.66E-18 |
| rs1107844 | T/C | 0.686 | 0.53 | 4.66E-18 |
| rs1107844 | T/C | 0.686 | 0.53 | 4.66E-18 |
| rs2079311 | C/T | 0.686 | 0.53 | 4.66E-18 |
| rs2079311 | C/T | 0.686 | 0.53 | 4.66E-18 |
| rs2079311 | C/T | 0.686 | 0.53 | 4.66E-18 |
| rs764218 | A/G | 0.686 | 0.53 | 4.66E-18 |
| rs764218 | A/G | 0.686 | 0.53 | 4.66E-18 |
| rs764218 | A/G | 0.686 | 0.53 | 4.66E-18 |
| rs764217 | G/C | 0.686 | 0.53 | 4.66E-18 |
| rs764217 | G/C | 0.686 | 0.53 | 4.66E-18 |
| rs764217 | G/C | 0.686 | 0.53 | 4.66E-18 |
| rs2412991 | T/C | 0.68 | 0.527 | 1.75E-17 |
| rs2412991 | T/C | 0.68 | 0.527 | 1.75E-17 |
| rs2412991 | T/C | 0.68 | 0.527 | 1.75E-17 |
| rs2267159 | C/T | 0.738 | 0.546 | 5.22E-17 |
| rs2267159 | C/T | 0.738 | 0.546 | 5.22E-17 |

| | | | | |
|---|---|---|---|---|
| rs2267159 | C/T | 0.738 | 0.546 | 5.22E-17 |
| rs5997676 | T/C | 0.699 | 0.501 | 3.53E-15 |
| rs5997676 | T/C | 0.699 | 0.501 | 3.53E-15 |
| rs5997676 | T/C | 0.699 | 0.501 | 3.53E-15 |
| rs5753187 | T/A | 0.699 | 0.501 | 3.53E-15 |
| rs5753187 | T/A | 0.699 | 0.501 | 3.53E-15 |
| rs5753187 | T/A | 0.699 | 0.501 | 3.53E-15 |
| rs2097871 | A/G | 0.703 | 0.494 | 5.35E-15 |
| rs2097871 | A/G | 0.703 | 0.494 | 5.35E-15 |
| rs2097871 | A/G | 0.703 | 0.494 | 5.35E-15 |
| rs4820863 | A/C | 0.643 | 0.465 | 3.00E-14 |
| rs4820863 | A/C | 0.643 | 0.465 | 3.00E-14 |
| rs4820863 | A/C | 0.643 | 0.465 | 3.00E-14 |
| rs5749122 | A/G | 0.582 | 0.404 | 1.33E-12 |
| rs5749122 | A/G | 0.582 | 0.404 | 1.33E-12 |
| rs5749122 | A/G | 0.582 | 0.404 | 1.33E-12 |
| rs11704977 | C/T | 0.193 | -0.529 | 3.54E-12 |
| rs11704977 | C/T | 0.193 | -0.529 | 3.54E-12 |
| rs11704977 | C/T | 0.193 | -0.529 | 3.54E-12 |
| rs5753191 | G/A | 0.556 | 0.407 | 7.57E-12 |
| rs5753191 | G/A | 0.556 | 0.407 | 7.57E-12 |
| rs5753191 | G/A | 0.556 | 0.407 | 7.57E-12 |
| rs5753192 | C/A | 0.518 | 0.385 | 1.64E-11 |
| rs5753192 | C/A | 0.518 | 0.385 | 1.64E-11 |
| rs5753192 | C/A | 0.518 | 0.385 | 1.64E-11 |
| rs60202446 | G/C | 0.22 | -0.466 | 2.09E-11 |
| rs60202446 | G/C | 0.22 | -0.466 | 2.09E-11 |
| rs60202446 | G/C | 0.22 | -0.466 | 2.09E-11 |
| rs5997677 | G/T | 0.571 | 0.4 | 2.12E-11 |
| rs5997677 | G/T | 0.571 | 0.4 | 2.12E-11 |
| rs5997677 | G/T | 0.571 | 0.4 | 2.12E-11 |
| rs6518702 | C/T | 0.487 | 0.371 | 7.86E-11 |
| rs6518702 | C/T | 0.487 | 0.371 | 7.86E-11 |
| rs6518702 | C/T | 0.487 | 0.371 | 7.86E-11 |
| rs11702947 | T/G | 0.198 | -0.485 | 1.43E-10 |

| | | | | |
|---|---|---|---|---|
| rs11702947 | T/G | 0.198 | -0.485 | 1.43E-10 |
| rs11702947 | T/G | 0.198 | -0.485 | 1.43E-10 |
| rs2267160 | C/T | 0.494 | 0.315 | 3.17E-08 |
| rs2267160 | C/T | 0.494 | 0.315 | 3.17E-08 |
| rs2267160 | C/T | 0.494 | 0.315 | 3.17E-08 |
